# Modelling optimal vaccination strategy for SARS-CoV-2 in the UK

**DOI:** 10.1101/2020.09.22.20194183

**Authors:** Sam Moore, Edward M. Hill, Louise Dyson, Michael J. Tildesley, Matt J. Keeling

## Abstract

The COVID-19 outbreak has highlighted our vulnerability to novel infections. Faced with this threat and no effective treatment, in line with many other countries, the UK adopted enforced social distancing (lockdown) to reduce transmission– successfully reducing the reproductive number *R* below one. However, given the large pool of susceptible individuals that remain, complete relaxation of controls is likely to generate a substantial second wave. Vaccination remains the only foreseeable means of both containing the infection and returning to normal interactions and behaviour. Here, we consider the optimal targeting of vaccination within the UK, with the aim of minimising future deaths or quality adjusted life year (QALY) losses. We show that, for a range of assumptions on the action and efficacy of the vaccine, targeting older age groups first is optimal and can avoid a second wave if the vaccine prevents transmission as well as disease.

## Introduction

After its initial detection in late 2019, the SARS-CoV-2 virus has spread across the globe with more than 20 million cases detected by mid-2020 [1]. For many individuals, infection develops into COVID-19 disease, with symptoms including fever, shortness of breath and altered sense of taste and smell, potentially escalating to a more severe state which may include pneumonia, sepsis, and kidney failure [2]. In general these symptoms and their severity have been observed to increase with age, such that it is the elderly that experience the greatest burden of disease. Due to the novelty of the virus, rapidity of its spread and the lack of effective treatments, mortalities (again predominantly in the older age-groups) have also been considerable.

By August 2020, six months after the initial cases in the UK, the country has reported in excess of 314,000 cases and more than 40,000 COVID-19 deaths [1]. As in many other countries, the virus spread in the UK has been mitigated by the introduction of a range of social distancing measures including the closing of workplaces, schools, pubs and restaurants, and the restriction of a range of leisure activities. Due to the continued risk of a considerable second wave, combined with the negative social and economic impact of continued social-distancing measures, a vaccine is urgently sought that may help curtail the global pandemic.

Mathematical models of COVID-19 dynamics and the impact of vaccination are important for public health planning and resource allocation. However, a substantial challenge of modelling vaccination against SARS-CoV-2 at this stage is the large number of unknown factors. At the time of writing, the UK has reportedly made deals for six different vaccine candidates created via differing approaches [3]: the Oxford (ChAdOx1 nCoV-19) [4, 5] and Janssen (Ad26.COV2.S) [6] vaccines, made from a genetically engineered virus; a vaccine developed by BioNtech/Pfizer (BNT162b1) [7], which uses an novel approach of injecting part of the virus’ genetic code; a vaccine created by Valneva (VLA2001) [8], which uses an inactive version of the virus; a vaccine created by Novavax (NVX-CoV2373) [9] and one under development by GlaxoSmithKline/Sanofi Pasteur [10], both using protein adjuvants to stimulate an immune response.

With an array of possibilities, differing in mechanism and still in varying stages of development, estimating response characteristics is infeasible. While the *ideal* vaccine would work to decrease susceptibility [11], thus limiting viral spread, the first successful candidate may just limit the occurrence or severity of symptoms. Moreover, it is still unclear when any vaccine will reach the final stage of being ready for mass deployment. Finally, while some promise has been shown in small human and animal studies - the success of the Oxford vaccine in a trial involving Rhesus Macaques [7] generated considerable excitement for instance - robust evidence as to how any vaccine candidate will perform in the wider human population is currently lacking [12].

Due to this uncertainty, we utilise a previously developed SARS-CoV-2 transmission model [13, 13] to understand the likely dynamics following the deployment of a vaccine. Rather than testing a single specific vaccine product, we assess three types of vaccines throughout our analysis to produce an evaluation that is as informative as possible for any likely set of vaccine characteristics. Explicitly, these comprise of: a vaccine that reduces susceptibility, thus being effective in inhibiting viral transmission as well as protecting the individual; one that reduces the probability of becoming symptomatic, which still has some benefit in reducing transmission as the model implicitly assumes that transmission from asymptomatic infections is less than from symptomatic ones; and one that protects against symptoms becoming severe, providing the direct protection against disease to the vaccinated individual only.

For each vaccine type we predict the impact of a range of possible scenarios by analysing sensitivity to vaccine efficacy, which may be uniform or age dependent, as well as the scale and targeting of deployment. Further, by considering different orders of prioritisation in terms of age group and health conditions, we arrive at an optimal targeting strategy for vaccination that achieves the greatest reduction in disease impact for the number of doses administered.

For a variety of other diseases, there is a precedent for combining modelling approaches with health economic evaluations to inform vaccine policy decisions based on a willingness to pay for each Quality Adjusted Life Year (QALY) saved [14-17]. Utilising this framework, we also consider how vaccination may be optimised to minimise the loss in QALYs, rather than simply the number of deaths. This methodology could allow a monetary value to be assigned to each dose of vaccine; while some cost benefit analysis has been pursued in relation to SARS-CoV-2 [18, 19], the unprecedented scale of the pandemic invalidates the usual metrics in this approach. We therefore do not utilise this health-economic approach as the main focus of our paper.

Despite the obvious need for both country-specific and more generic forecasts of the effects of vaccination on the COVID-19 outbreak, it has received relatively limited attention. As far as we are aware only two studies exist, both based on US data [20, 21]. Our study adds a UK perspective to these studies and generates a wider perspective on how the type of protection offered by the vaccine as well as the underlying efficacy interact with targeting risk-groups to generate maximal benefit.

## Model Formulation

We used a compartmental age-structured model, developed to simulate the spread of SARS-CoV-2 within regions of the UK [22], with parameters inferred to generate a good match to deaths, hospitalisations, hospital occupancy and serological testing [13]. It involves an extended SEIR-type framework: susceptibles (S) may become infected and move into a latent exposed (E) state before progressing to become infectious. Echoing the observed behaviour of COVID-19 infections, the model differentiates between individuals who are symptomatic (D, likely to be detected) and those who are asymptomatic (U, likely to remain undetected). Partitioning those infectious by symptom status allows for the lower level of transmission believed to be associated with asymptomatic infection. It also generates the possible progression of symptoms increasing in severity, leading to hospitalisation and/or death (Fig 1).

**Fig. 1:**
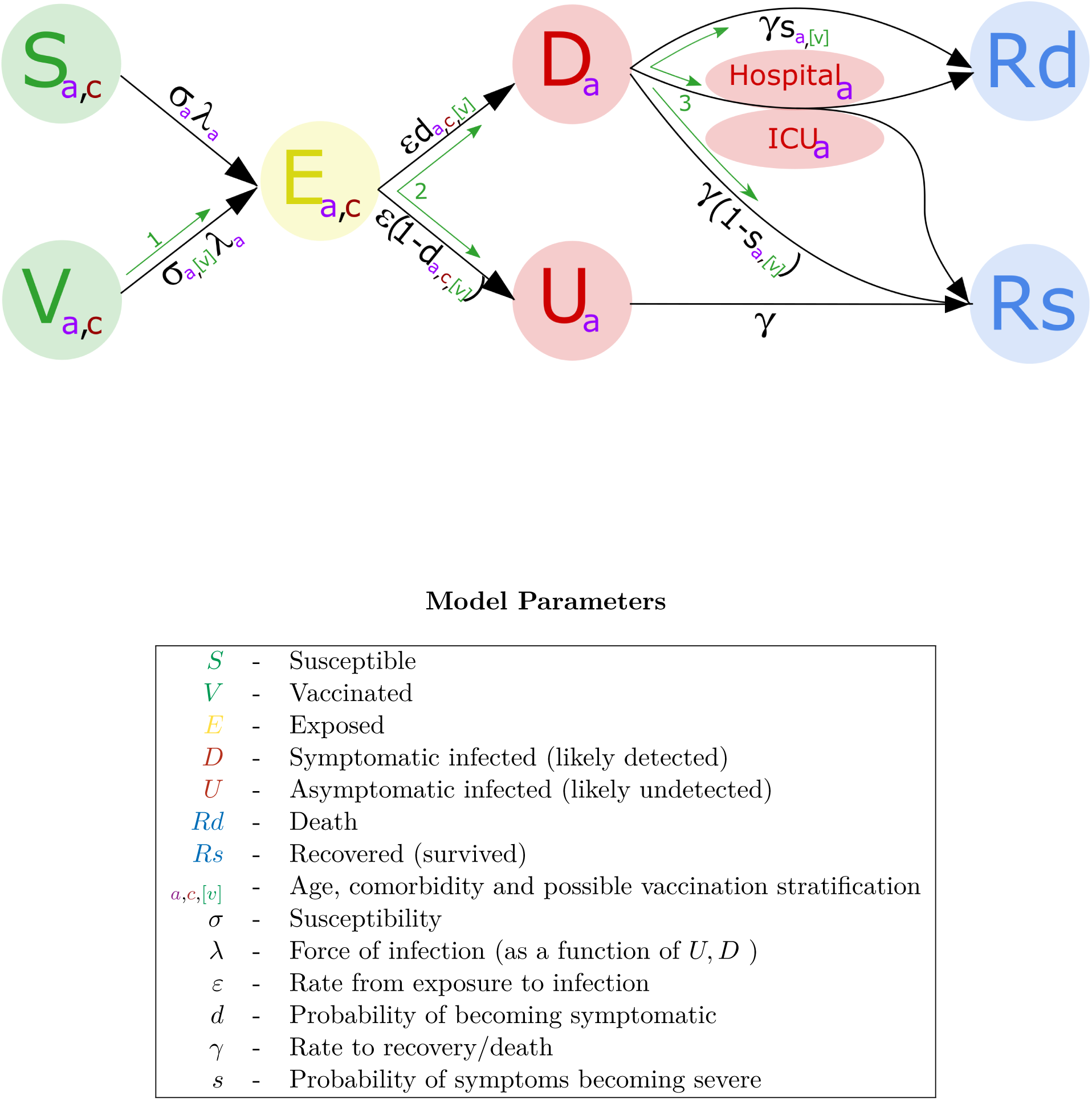
Representation of model states and transitions. The three different possible types of vaccination are shown by green arrows and bracketed variables. These are: 1. Reduction in susceptibility. 2. Reduction in becoming symptomatic. 3. Reduction in experiencing severe symptoms.

The model is stratified by age structure, with force of infection determined by the use of age-dependent (who acquires infection from whom) social contact matrices for the UK [23, 24]. Additionally, we assumed susceptibility and the probabilities of becoming symptomatic, being hospitalised and dying to be age dependent, and are matched to UK outbreak data. Finally, our model formulation accounted for the role of household isolation by allowing *first* infections within a household to cause new *secondary* infections at an increased rate (more details may be found in [22]). This allows secondary household contacts to be isolated and consequently play no further role in the outbreak. Model parameters were inferred on a regional basis using local time series of recorded daily hospitalisation numbers, hospital bed occupancy, ICU occupancy and daily deaths [13].

This model formulation is then extended to capture a range of vaccination scenarios. Below we detail how vaccination of different forms is introduced into the model; the degree to which non-pharmaceutical interventions are reduced; and how the population is partitioned according to high-risk comorbidities - to assess if these are a priority group for immunisation.

### Comorbidity

Alongside age, it is well understood that underlying health conditions are critically important in determining the likelihood of individuals to experience severe symptoms; in younger, healthier populations a relatively low symptomatic rate has been observed [25-28] and there are few severe cases. To account for heterogeneity of risk that is not attributable to age, we therefore allowed the probability of experiencing severe health outcomes (as a result of COVID-19 disease) to be dependent on underlying health conditions.

We calculated the additional risk by comparing the prevalence of conditions amongst COVID-19 mortalities within the general population. For the purpose of simplification, we used a binary system to divide the population into those with significant health conditions and those without, rather than considering individual conditions separately [29].

We estimated the proportion of the population at heightened risk, due to the presence of a cormobidity, using the twelve health conditions with the highest associated risk factors as identified in a recent study by Williamson *et al*. [30]. We summarise these conditions, alongside their prevalence and individual risk, in table 1.

For the 18.42% of the population with one or more of the comorbidity conditions listed in Table 1, we calculated that the average increase in the risk of morbidity is 2.43 (mean value) - suggesting that individuals with these risk factors are more than twice as likely to die from COVID-19 infection compared to others of the same age. The distribution of these combined symptoms by age group was calculated using data from the Royal College of General Practitioners Research and Surveillance Centre [31] combined with statistics on cancer and diabetes prevalence found in [32, 33].

The comorbidities associated with increased risk from COVID-19 infection are found to occur predominantly in elder age groups (Fig 2a), with individuals above 75 years of age more likely than not to have underlying health problems. When these factors are incorporated into the predictive model, we see that deaths are dominated by older age groups and individuals with underlying comorbidities (Fig 2b).

**Fig. 2:**
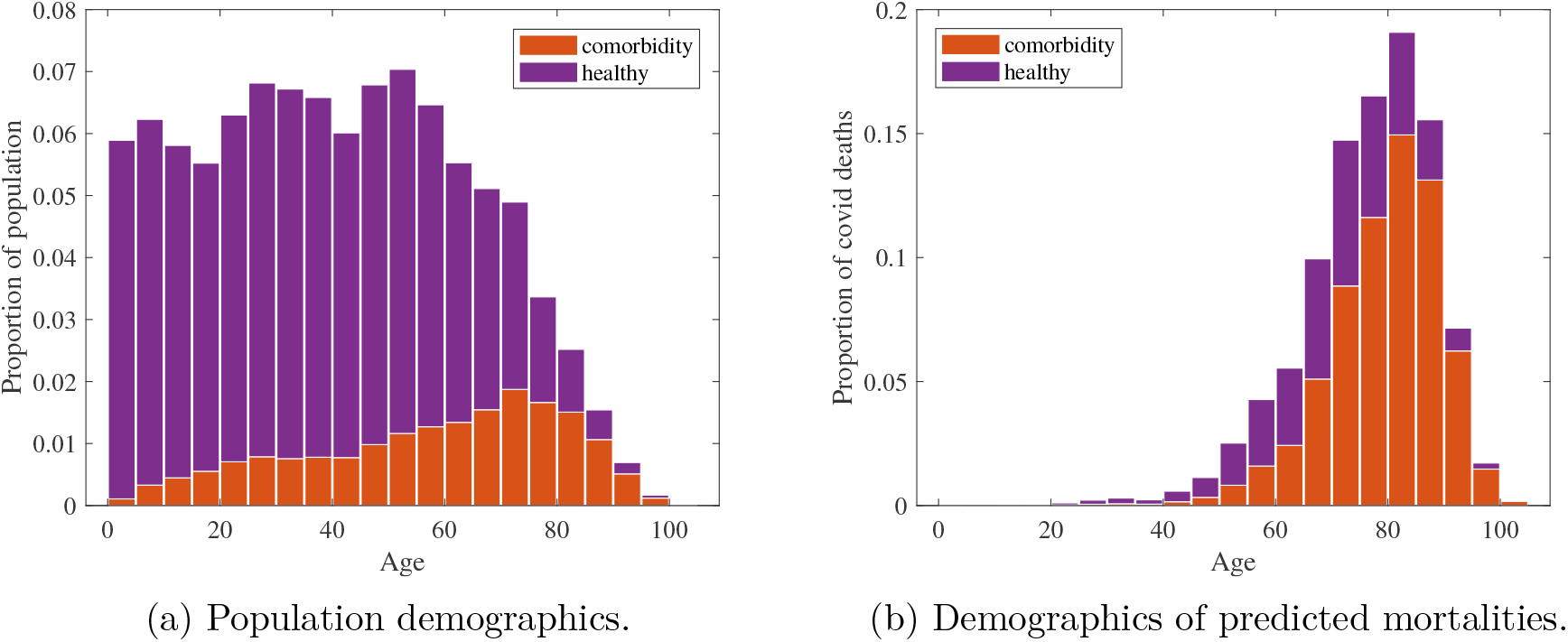
The percentage of (a) the population and (b) deaths predicted by our model, within each five-year age band. The proportions of each age group with health conditions are shown in red, with the proportion of each age group without health conditions (healthy) shown in purple.

**Table 1:** Table of identified significant health conditions as calculated in [30]. For each condition we give the estimated prevalence in the population as well as the increased risk of death found as a hazard ratio calculation adjusted for demographics and coexisting conditions.

| Condition | Percentage of population | Increased risk (CI) |
| --- | --- | --- |
| Diabetes | 9.9 | 2.06 (1.88-2.25) |
| Kidney disease | 6.3 | 1.72 (1.62-1.83) |
| Heart disease | 6.1 | 1.27 (1.20-1.35) |
| Respiratory (excl. asthma) | 4.1 | 1.78 (1.67-1.90) |
| Dementia/stroke | 2.1 | 1.79 (1.67-1.93) |
| Severe asthma<sup>1</sup> | 1.7 | 1.25 (1.08-1.44) |
| Immunosuppressive conditions | 1.6 | 1.69 (1.21-2.34) |
| Cancer<sup>2</sup> | 1.2 | 2.51 (2.01-3.15) |
| Neurological (excl. dementia/stroke) | 1.0 | 2.46 (2.19-2.76) |
| Liver disease | 0.7 | 1.61 (1.33-1.95) |
| Spleen diseases | 0.2 | 1.41 (0.93-2.12) |
| Organ transplant | 0.1 | 4.27 (3.20-5.70) |

<sup>1</sup> Severity defined by recent oral corticosteroid use.
<sup>2</sup> Limited to cases that have either been diagnosed in the last year or involve haematological malignancy.

### Social Measures

The use of non-pharmaceutical social-distancing (or lockdown) measures has already proven to have substantially reduced the scale of the pandemic [34]. We include such measures into our model through a reduction in the relevant contact matrices (i.e. scaling down matrices associated with schools, work places and other settings) whilst increasing the level of contact within household environments [13, 13]. Additionally, we incorporate household quarantining by transferring a proportion of the symptomatic individuals to a quarantined state with a still greater reduction in external contact. Test and trace is not yet included in the model, due to lack of data on its current scale or effectiveness.

It is expected that a sustained level of social-distancing measures will continue to be used to mitigate the disease until pharmaceutical approaches are available. As such, in all simulations, we initially simulate the first wave of the UK epidemic to date, followed by a continuation of current lock-down measures (generating *R ≈* 1.0) until vaccination is completed.

As exemplified by simple models [35, 36], the impact of any vaccination campaign is highly dependent upon both the proportion of the population successfully immunised and the reproductive ratio. As such, we expect the success of any COVID-19 vaccine programme to depend on the reproductive ratio *R* when the programme begins, which in turn is dependent upon the level of non-pharmaceutical interventions (NPIs) and the proportion of the population already infected. Fig 3 compares simulations in which all NPIs are lifted at the start of 2021 (in which case *R* ≈ 2.3), with a scenario in which a limited number of NPIs are still in place (leading to *R* ≈ 1.8). Given the general level of awareness and concern in the population, we take *R ≈* 1.8 as our base-case during the majority of our simulations, but do assess a return to pre-COVID behaviour (*R* ≈ 2.3) on completion of the vaccination campaign.

**Fig. 3:**
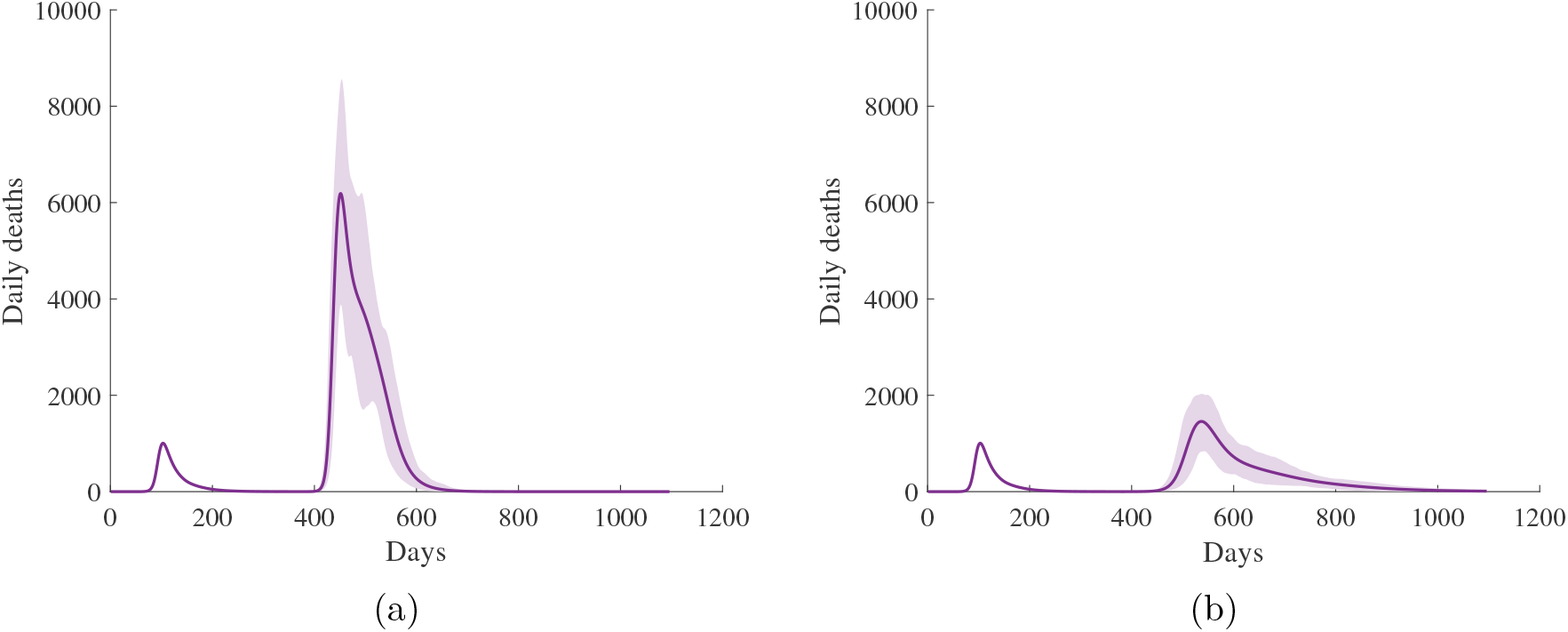
Projected daily deaths without vaccination given (a) no new control measures leading to *R =* 2.3(±0.2), and (b) limited control measures sufficient to reduce the basic reproductive number to approximately *R* = 1.8(±0.1). Uncertainty in numbers is represented by the shaded region, within which 95% of simulations are found to fall. Simulations do not include random reintroduction, which may cause a much higher level of uncertainty in the timing of the second outbreak.

### Vaccination

The novelty of the virus, combined with the urgency in finding a response, has resulted in a plethora of possible vaccine candidates in various stages of development across the globe [37, 38]. To account for uncertainty in both the type and the degree of protection a vaccine may provide, we test the effectsof three different vaccination mechanisms within our model, each over a range of efficacy:

1. Reduction in susceptibility (type 1): We reduce the rate at which the vaccinated susceptibles may become infected. This is the most effective form of vaccine as it may significantly reduce *R* as well as protecting the individual.
2. Reduction in becoming symptomatic (type 2): The age and health dependent probabilities of becoming symptomatic are adjusted according to vaccine efficacy. This type of vaccine therefore has a substantial impact on disease, but also has some impact on infection spread (and hence *R*) due to asymptomatic individuals being less infectious.
3. Reduction in experiencing severe symptoms (type 3): The age dependent probabilities of symptoms becoming severe (leading to hospitalisation and/or death) are adjusted according to vaccine efficacy. This type of vaccine only protects the individual and does not impact the transmission dynamics.

The action of these three vaccination types are shown in Fig 1. It should be noted that, whilst instantaneous vaccination may not be a literal possibility, due to the uncertainty on time scales combined with the potential for a second infection wave to be delayed by social-distancing measures during vaccine deployment, it is a reasonable modelling assumption for our exploratory purposes.

### Simulation specifics

In all simulations, we ran the model for an initial period (until the end of 2020) without any form of vaccination, to create a base-line population of susceptible, infectious and recovered (immune) individuals that approximates the state of population when vaccination begins. Given that during this period *R* is close to, but below one, the precise timing of the start of the vaccination programme has little effect. Similarly, if NPIs are maintained until vaccination is complete, the speed of vaccination is unlikely to play a major role.

At the start of 2021, to simulate rapid immunisation, we transfer a specified proportion of the remaining susceptible individuals into a vaccinated state; the action of infection on individuals in the vaccinated class is dependent on vaccine type.

We now outline the four/five different approaches we use to quantify the optimal targeting of vaccine and the sensitivity to key uncertainties.

### Priority order of vaccination

To determine a vaccination priority order for who should receive the vaccine first, we ranked each group by the ratio of the number of doses administered to the reduction in the outcome measure of interest. Our two selected outcome measures were: (i) deaths post-2020; (ii) quality adjusted life years (QALYs) lost post-2020. We divided the population into 20 year age bands (0-19,20-39,40-59,60-79,80+), together with an additional group of people with comorbidities independent of age. For each vaccination scenario considered, we exhaustively tested the order of deployment amongst these groups.

### Prioritisation of healthcare workers

Due to their significantly heightened exposure, another group that may benefit from early targeted vaccination is healthcare workers (HCWs). Proportionally, 11.6 times more cases have been observed in UK HCWs than the general populous - this can partially be explained by the increased testing in hospital environments. Yet, even accounting for this increased testing, HCWs are estimated to experience a 3.40 (3.37-3.43) fold increase in infection risk [39].

We included this increased risk to HCWs in our model by adjustment of the susceptibility parameters, using a similar methodology that was applied to underlying health conditions. This increased susceptibility was applied to all of the 1,134,824 full time NHS employees [40], which we assumed to be evenly distributed (age and region proportional) amongst the working age group (20-64).

### Sensitivity to vaccine characteristics

Using the identified optimal prioritisation order for the given vaccine type, with a maximum vaccine uptake of 70% across each sub-group in the population, we analyse the sensitivity of number of deaths and QALYs lost to vaccine efficacy, considering vaccines with 50%, 70% and 90% efficacy. Here we define an efficacy of *n*% to give an *n*% reduction in susceptibility/chance of becoming symptomatic/chance of developing severe symptoms for a type 1/2/3 vaccine respectively.

### Sensitivity to speed of vaccine deployment

In the majority of the modelling we assume that the time needed to vaccinate the population and for the vaccine to confer immunity is short. This assumption has little impact if the reproduction number *R* is below one while the vaccine program is being undertaken.

To explore the impact of slow vaccine deployment in the face of increasing infection we utilise simulations in which NPIs are relaxed, to bring *R* up to 1.8, two months before a type 1 vaccine with 70% efficacy begins deployment, thus delivery occurs alongside an already widespread and increasing epidemic. We simulate gradual delivery dynamics by running our model over daily time steps with fixed number of susceptible or asymptomatic individuals being successfully immunised at each interval. More realistic delivery scenarios may be simulated as vaccination deployment plans are better understood.

### Sensitivity to age-dependent efficacy

Many existing vaccines are not uniformly effective and response may vary significantly due to factors including age [41]. In the case of a virus where severity of symptoms are age-dependent, like SARS-CoV-2, age-dependent vaccine efficacy may have a significant impact. Of particular concern is whether an age-dependent decline in efficacy could alter the group priority order for receiving a vaccine, and the potential scale of the subsequent outbreak. We test the likely implications of reduced vaccine effectiveness in the elderly by considering an array of different efficacy profiles consisting of a base efficacy uniform amongst those below the age of 45 with linearly declining efficacy to a minimal value shared by those above the age of 85.

For each of the scenarios described above and for each combination of considered vaccine type, efficacy and priority order, we performed 100 separate simulations. Each simulation used a distinct parameter set drawn from posterior parameter distributions, which were obtained from the inference procedure that tuned the model to the available COVID-19 health outcome data streams for the UK [13].

## Results

We now consider the ordering of vaccination and its impact in terms of the number of deaths and associated QALYs lost. For the majority of the situations we consider, we plot the number of individuals vaccinated on the x-axis (assuming a maximum of 70% of susceptible or asymptomatically infected individuals are vaccinated in any group) and the resultant number of deaths or QALY losses on the y-axis. The priority ordering is determined by which groups generate the greatest reduction for the number of doses administered (steepest decline on the graphs).

In every case, optimising for either a reduction in death or a reduction in QALYs was found to give consistent ordering, due to the substantive contribution of mortality events to the overall QALY loss (Fig 4). When structuring by age alone, the most efficacious reduction was found through an oldest first approach - despite not being the most crucial group in terms of transmission, the considerably heightened vulnerability amongst the elderly means that priority should be given to protecting them directly. We also highlight the substantial advantages of a strategically targeted vaccine over an unbiased (random) approach in every scenario (blue line, Fig 4).

**Fig. 4:**
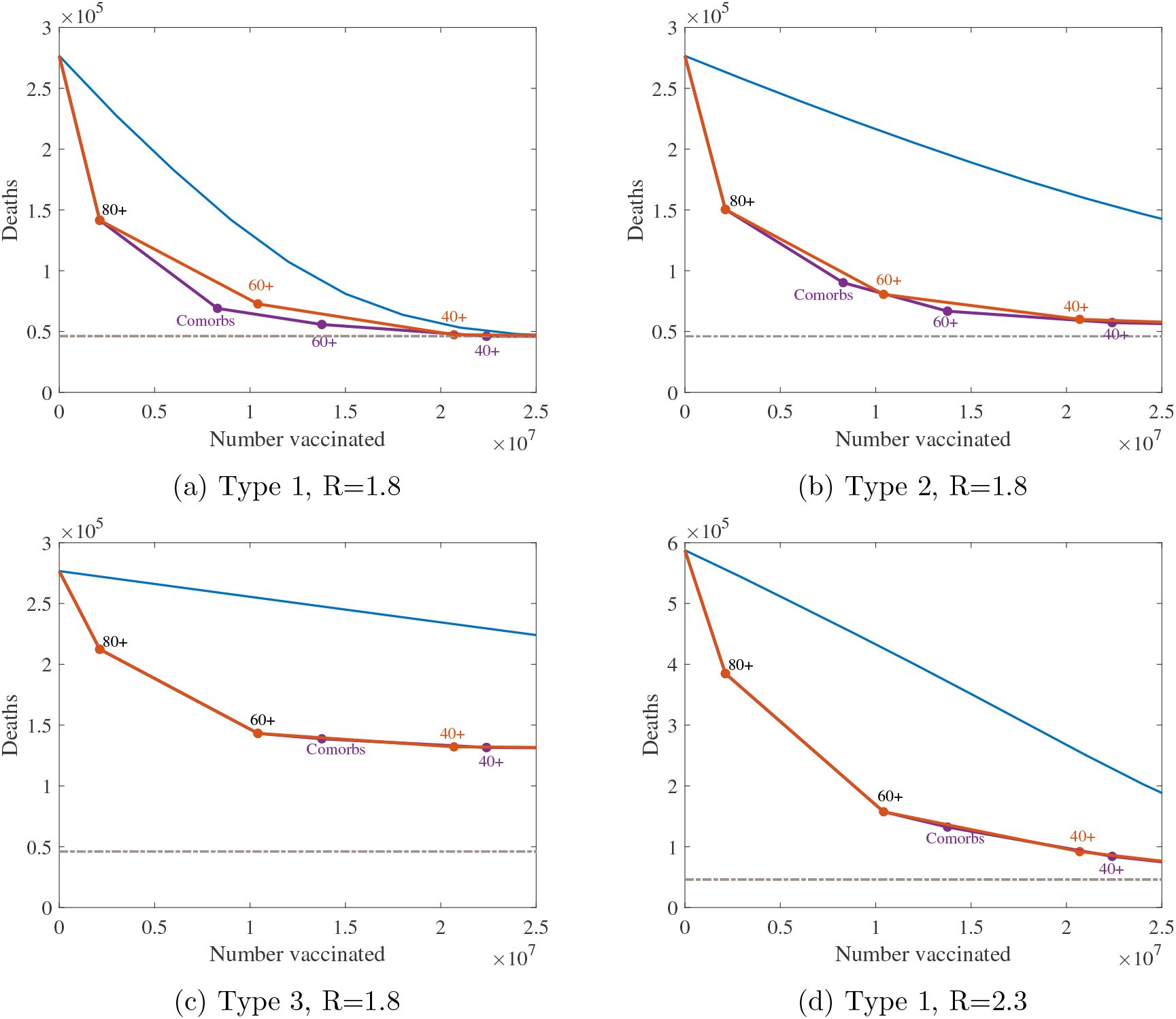
Comparison of vaccination ordering for four different vaccination scenarios. In each panel, the purple lines show the optimum vaccination ordering of groups comprising of 20 year age bands together with comorbities across all ages, and the orange line for age groupings only. The optimum strategy is identified by giving greatest reduction in deaths per vaccination in each instance. The blue lines show an unbiased strategy with vaccinated numbers dispersed evenly across the population and the grey dotted line show the base level of morbidity from the first infection wave.

In contrast, there was less consistency in the optimal position of vaccinating comorbidities in the priority order, which varied between just before and just after the 60-80 age group. One such instance in which the discrepancy may be seen is when contrasting the type 1 and type 3 vaccine with a low level of NPIs (*R* ≈ 1.8, Fig 4 a and b). While the cost-benefit implications of this variation are often found to be small, due to the large overlap between the group of people with comorbidities and the elderly, it is substantial enough that it may be worth critical consideration in a scenario where vaccine quantity and/or deployment rate are limited.

### Healthcare workers

We now consider the addition of healthcare workers (HCW) as an additional risk group, and assess their optimal position in the order of vaccination priority. For a type 1 vaccine (for which there is benefit in targeting infection spreaders as well as the vulnerable) we associate high important to the vaccination of HCW, second only to the most elderly age group of those aged 80 and above (Fig 5). It is important to note that our treatment of HCWs only accounts for increased personal risk. We have not taken into account the increased contact between HCWs and the vulnerable which may significantly amplify their risk as spreaders of infection, increasing their priority for vaccination. Given also the relatively small number of HCWs (less than 2% of the population), we conclude that this group should be included as a high priority group for vaccination. However, for vaccines of type 2 or 3 that do not significantly reduce infection spread, the HCW group may be considerably less important and their priority should be judged accordingly.

**Fig. 5:**
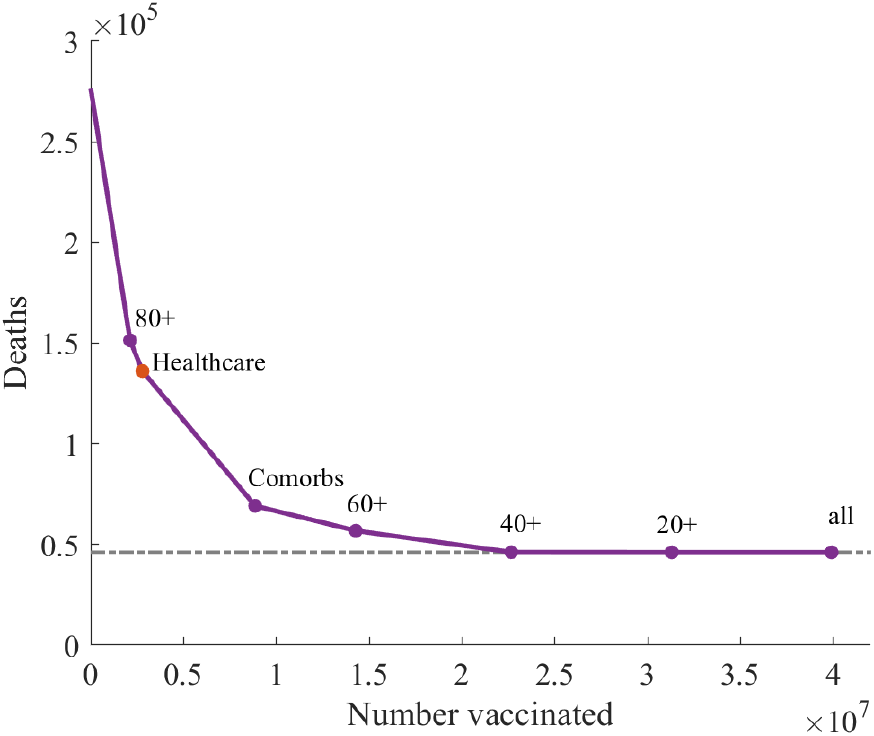
Optimal vaccination ordering for age, comorbidity and HCW groups in a scenario with a type 1 vaccination and low level social measures (*R* = 1.8).

### Vaccine type comparison under optimal priority order

For the three types of vaccine action, and for three different levels of efficacy we consider the impact of a vaccine program targeted by age and comorbidity under weak non-pharmaceutical interventions (*R* ≈ 1.8). For Type 1 vaccines, which have the greatest impact, we also consider complete lifting of all non-pharmaceutical interventions (*R* ≈ 2.3).

### Type 1 vaccine

Due to the ability of a type 1 vaccine in preventing the spread of infection as well as protecting specific individuals, we find that such a vaccine, even with relatively low efficacy, could be highly effective in preventing further COVID-19 mortality when combined with limited social-distancing measures (i.e when *R* ≈ 1.8).

The best performing prioritisation order begins with those aged 80 and above, followed by those with health conditions, before the rest of the population in age order. Under such an ordering, we estimate a vaccine with 50% efficacy delivered to 70% of the population above the age of 20 to be sufficient to prevent a SARS-CoV-2 resurgence in our considered scenario (Fig 6 top row), although a more efficacious vaccine could achieve the same results with only vaccinating those above 40.

**Fig. 6:**
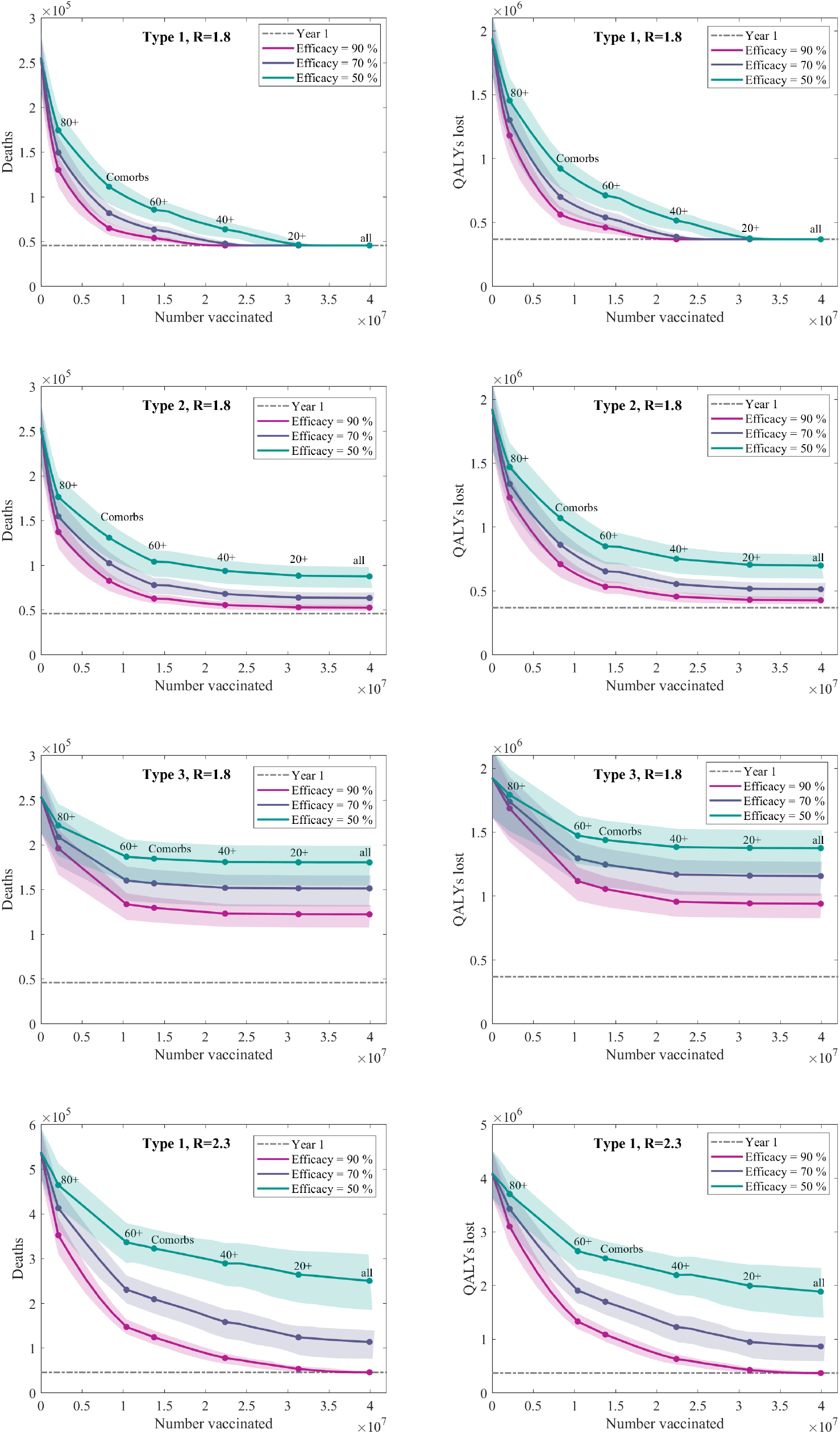
Deaths and QALYs lost versus the proportion of the population vaccinated. We use the optimal vaccination ordering with a maximum vaccine uptake of 70% across the population. We display the mean values of 100 independent simulations for each vaccine efficacy (solid lines), with 95% prediction intervals (shaded regions). The grey dotted line shows the base level of mortality from the first wave of the pandemic.

### Type 2 vaccine

When combined with limited social-distancing measures (such that *R* = 1.8), a type 2 vaccine (that reduces the chance of becoming symptomatic) with reasonable efficacy may still be sufficient to prevent a significant mortality in a second wave (Fig 6 second row). However, due to only providing a limited reduction in transmission little additional benefit is to be found in vaccinating those in low risk (younger age) categories. Again we identify that it is best to prioritise those above the age of 80, followed by those with health conditions, before the rest of the population in age order. For type 2 vaccines that generate an efficacy of 50% or below we would predict a large-scale second wave of infection when control measures are relaxed with a comparable number of deaths to experienced in the first wave.

### Type 3 vaccine

A type 3 vaccination, which only reduces the chance of a recipient experiencing severe disease, does not reduce the risk of transmission and therefore we predict a large scale second outbreak when containment measures are lifted, comparable to that shown in Fig 3b. However, it can still generate considerable benefit when deployed to high risk individuals, particularly those over the age of 60 (Fig 6 third row). For the most efficacious vaccines, immunising 70% of those over 60 years old could potentially halve the number of deaths in the second wave.

Contrary to the other three vaccination types, here we find it best to prioritise 60-80 year olds above all those with health conditions. We postulate this is due to the lack of reduction in transmission, such that the higher degree of social contact amongst the younger people within the comorbidity group is not an influencing factor.

### Type 1 vaccine, without measures

The most desirable vaccine would be one that is sufficient to entirely contain the pandemic without the need for any other intervention (when we estimate *R ≈* 2.3). Amongst our set of vaccine scenarios, we find the only plausible candidate for preventing a SARS-CoV-2 resurgence (in the absence of any other intervention) would be a type 1 vaccine with an efficacy above 80% and with 70% of the entire population vaccinated (Fig 6 bottom row). Other levels of vaccine efficacy would naturally trade-off with a different proportions of the population needing to be vaccinated; similarly a higher uptake in the older at-risk populations would also mean that fewer younger individuals would need to be immunised.

For a type 1 vaccine in a scenario without additional measures (*R ≈* 2.3), we find scant difference between taking a fully age ordered strategy, and one including comorbidity, leaving no clear point at which those we categorise as having health conditions should be prioritised (Fig 4d).

### Slow deployment

The results presented so far are based on a model with instantaneous vaccination delivery. This is deemed sufficient for a generic understanding of the impacts of vaccination, varying uptake levels and targeting. Additionally, if NPI remain in place (such that *R* < 1) until vaccination is completed, the time taken to implement a slower vaccination campaign will lead to there being fewer cases when NPIs are finally relaxed. It is also the case that, since temporal factors such as deployment speed and a start date for delivery are largely unknown, there is limited value in incorporating complex dynamics at this time. More realistic delivery scenarios may be simulated as vaccination deployment plans are better understood.

In a scenario where *R* is greater than one during the vaccine program, such that there is a race between vaccination to reach herd-immunity and the rise of the epidemic, the speed of vaccine deployment is key (Fig 7); a rapidly deployed untargeted campaign is far more successful than a slow but optimally targeted one. Importantly, we find that the optimal vaccination strategy (in terms of the ordering of age and risk groups) is independent of delivery speed. However, for very rapid vaccine deployment (when all groups can be vaccinated before there are many more infections) or for very slow deployment (when the epidemic is complete before many individuals are immunised) the ordering is far less important.

**Fig. 7:**
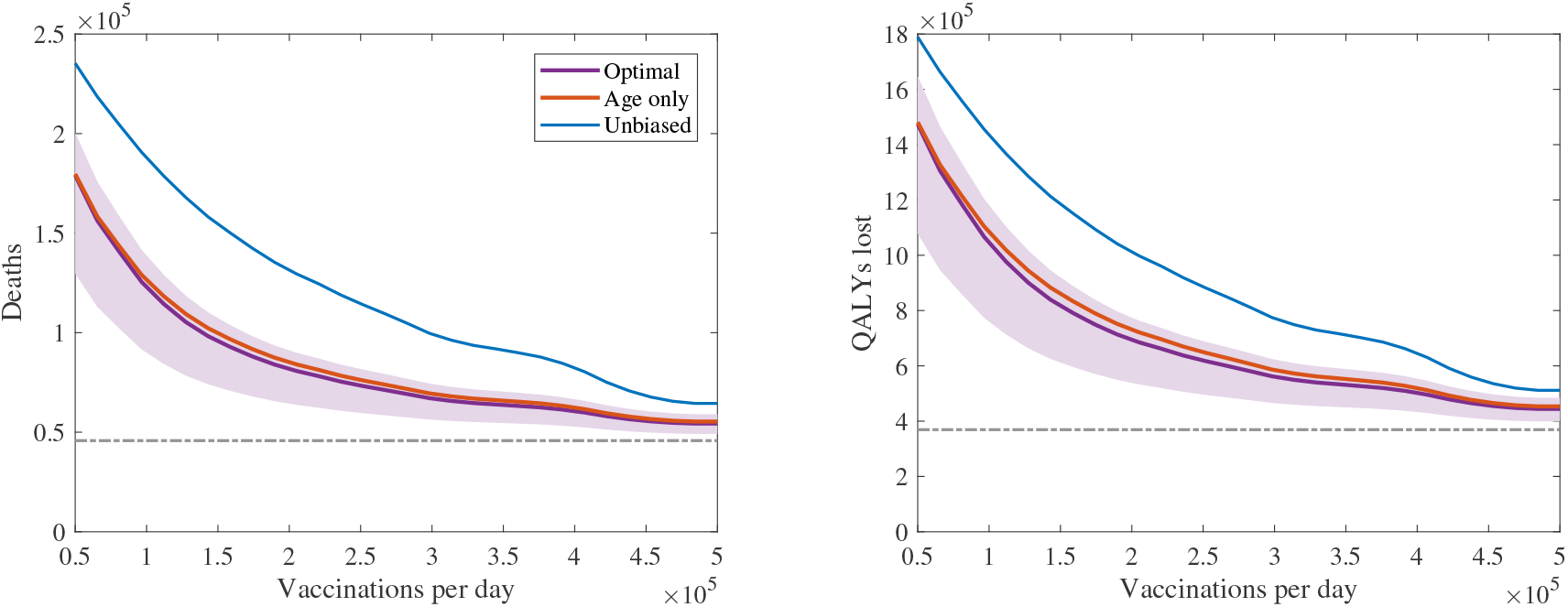
The total number of (left) deaths and (right) QALYs lost resulting from a second epidemic wave vs speed of deployment for a type 1 vaccination with 70% uptake and 70% efficacy. Vaccine deployment is started 2 months after stricter NPIs are relaxed to leave low level measures sufficient to keep *R ≈* 1.8. The purple line (and shaded prediction region) represents projected outcomes under the identified optimal ordering with age and comorbidity groups, the orange line outcomes using the identified optimal ordering accounting for age groups only (without comorbidity), and the blue line outcomes with an unbiased population wide delivery.

### Age-dependent efficacy

To simulate reduced efficacy of the vaccine in older age groups, we allow efficacy to be age-dependent - attaining a high maximum value for individuals below age 45, then dropping linearly to age 85 when the efficacy reaches a minimum value (Fig 8a inset).

**Fig. 8:**
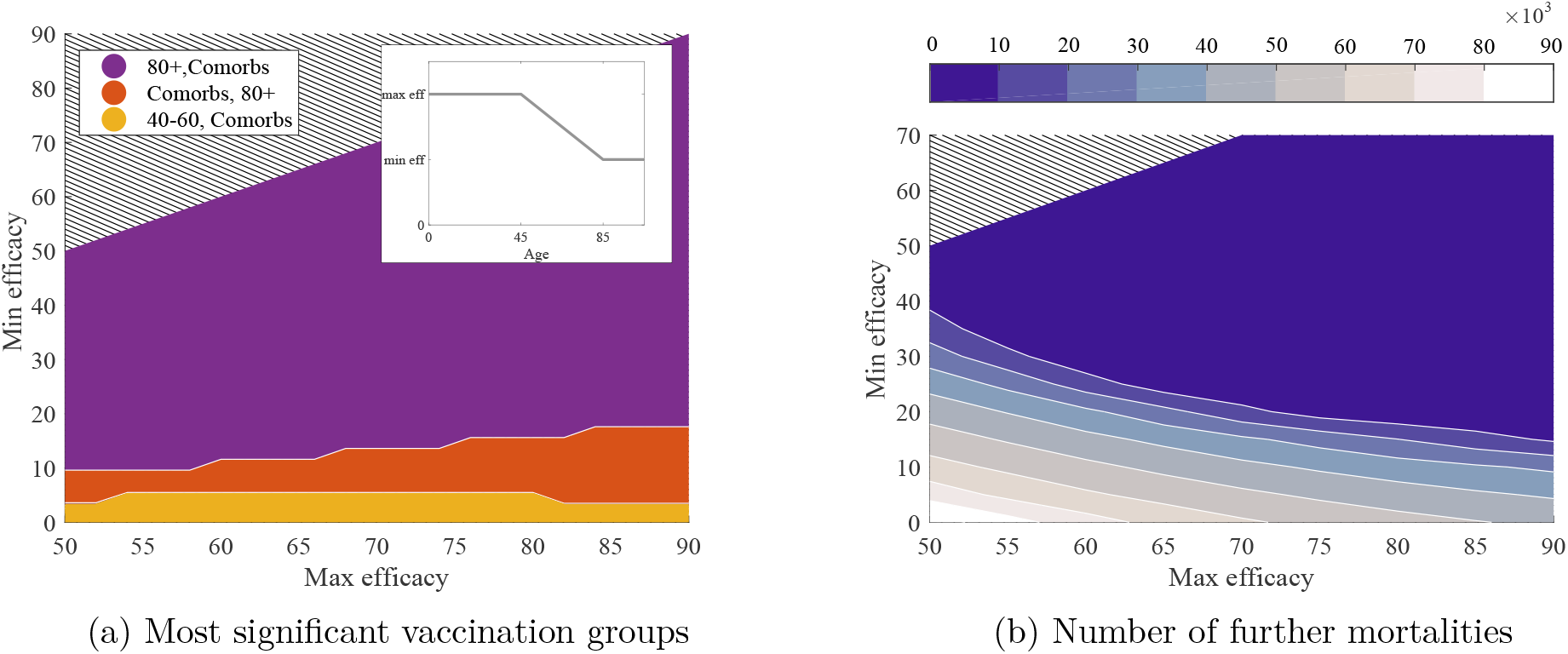
The effect of different levels of declining type 1 vaccine efficacy (*R* ≈ 1.8) on both the optimal vaccine ordering and success at limiting mortality. A maximum efficacy value is applied to all individuals below the age of 45 and a minimum level to individuals above the age of 85. Efficacy is assumed to decay linearly between these two levels to give the efficacy for intermediary age groups. This distribution is represented by the inset in panel (a). Panel (a) shows when, dependent on minimum and maximum efficacy, the two groups deemed most significant for vaccination impact vary. In the purple region it is optimal to vaccinate those above the age of 80 followed by comorbidities, in the red comorbidities followed by 80+ and in the yellow those in the 40-60 age group followed by comorbidities. Panel (b) shows the expected further mortality resulting from a second infection wave with 70% of the whole population vaccinated for different minimum/maximum type 1 vaccine efficacies. The large dark blue region corresponds to less than 10,000 deaths in any second wave following vaccination.

There is again a surprising robustness in ordering (Fig 8a) with the elderly (over 80 years old) remaining the key group to initially target, even when the efficacy in this age group drops to just 20%. Even lower efficacies necessitate a switch with individuals with underlying comorbidities or aged 40-60 being prioritised. Lower efficacy vaccines are, unsurprisingly, associated with larger subsequent epidemics and therefore more deaths; much of this increase is concentrated, however, in low efficacy regions of parameter-space, with the estimated number of deaths being relatively constant across a range of efficacy values for both older and younger age-groups.

While here we focus on a type 1 vaccine; the results for vaccine types 2 and 3 are found to be more extreme, as the drop in efficacy aligns with the age-groups most in need of protection. For a type 3 vaccine the elderly are found to remain the initial targeting priority up to a reduction in efficacy down to just 10%.50

## Discussion

It has often been stated that the only way for population behaviour to return to normal is through achieving herd-immunity. Allowing this to occur through natural infection is far too dangerous, even if the most at-risk groups are shielded, making vaccination the only viable alternative. While there are huge efforts being undertaken to develop a vaccine, less attention has been paid to how such a vaccine would be deployed. For SARS-CoV-2, the infection is relatively homogeneous across age-groups with a slight bias towards younger adults [42], but disease - especially severe disease - is associated with old age and a range of comorbidities. As with many vaccines, there is tension between vaccination of those most responsible for driving transmission (vaccination to reduce *R*) and vaccination of those most likely to suffer severe health outcomes (vaccination to limit disease) [43]. Here, using detailed mathematical models that have been matched to UK COVID-19 outbreak data, we find that vaccine strategies targeting the elderly are optimal in terms of reducing future mortality (Fig 4), even if vaccinating younger group-ages would have a greater impact on the reproductive number, *R*.

We have found that five main factors influence the success of any vaccination programme (Fig 6).

1. The characteristics of the vaccine: vaccines that reduce susceptiblity and therefore prevent onwards transsmission (type 1 vaccines) lead to a far greater reduction in mortality, compared to vaccines that reduce disease (types 2 and 3);
2. The efficacy of the vaccine: vaccines of all types with higher efficacy generate both greater protection of individuals and (for type 1 and 2 vaccines) greater protection within the population;
3. The reproduction number once the vaccination program is complete: higher reproduction numbers require a greater proportion of the population to be immunised to prevent a second outbreak (mimicking the classic *V_c_* = 1 − 1/*R*_0_ paradigm). If all non-pharmaceutical interventions (NPIs) are lifted, and if approximately 70% of the population are vaccinated, only highly efficacious, type 1 vaccines are able to control subsequent waves of infection;
4. The proportion of the population vaccinated: throughout we have assumed that at most 70% of any age or risk group would be vaccinated, however there is a natural relationship between vaccine efficacy and proportion vaccinated that generates the same level of overall protection;
5. Who is vaccinated: we have consistently shown that prioritising the vaccination of the elderly is by far the most effective strategy for reducing the number of deaths in any second wave. It is only for vaccines that have a vastly reduced efficacy in the elderly (below 20%) that other prioritisation orderings become more effective.

In the majority of this work, we have focused on the resultant outbreak after protection by vaccination is complete. It is likely that any future vaccine will require (at least) two doses, and there will be a delay between vaccination and protection. If the reproductive ratio remains below one while these events take place, then in general the delays are irrelevant. However, in the scenario where NPIs are relaxed before immunisation of the population is complete, rapid deployment of the vaccine is critical to success (Fig 7).

Our analysis has also focused throughout on two differing outcome measures, both the number of deaths and the expected QALY loss. Given that QALYs give greater weight to preventing illness or death in younger age-groups, it is somewhat surprising that the optimal order of vaccination remains unchanged. This can only be attributed to the far greater severity of disease experienced by the elderly. We have purposefully not performed a full cost-benefit analysis on the vaccine, as in many ways the outcome of this pandemic is difficult to capture in monetary terms. However, under the assumption that *R ≈* 1.8, an unvaccinated population would experience a loss of 1.58 (1.29-1.78) million QALYs during a second wave, which could be prevented by vaccination of 20.6 (18.9-22.3) million targeted individuals or 41.2 (37.8-44.6) million doses of a vaccine with 90% efficacy (assuming the need for a primary and booster dose). This simple calculation, together with a standard cost-effectiveness threshold of £20,000 per QALY, would suggest that the UK should be willing to pay around £767 (£578-£942) per dose of vaccine - highlighting its huge public health benefit.

Our mathematical model has been carefully matched to the observed dynamics in the UK [13], capturing the fundamental age-structured epidemiology. In contrast, little is known about the characteristics of any new vaccine and the associated model parameters. Many more details could therefore be incorporated within the model structure when these become known for any emerging vaccine candidate. We have, for simplicity, assumed 70% vaccine uptake across all age-groups based on what has been obtainable for other vaccines, such as within elder age groups and healthcare workers for the UK seasonal influenza vaccination programme [44]. It is likely that the public’s response to a SARS-CoV-2 vaccine may be different, depending on perceived risk which is likely to be age dependent. Similarly, the assumed efficacy has either been taken as constant across all age-groups or obeying a simple declining function with age; incorporating more realistic estimates about the action of any vaccine is a vital first step in assessing the benefits. To a large extent, we have also ignored the time delays between the start of any vaccine program and the protection of the vaccinated individuals; the full implications of any delays can only be calculated once more is known about the mechanisms and limits on delivery, as well as a better understanding of the reproductive number (R) during and after the campaign. Including these elements within the model is feasible, but is presently hampered by a lack of data for parameter inference.

This work has focused on the dynamics within the UK, but the robustness of our conclusions, that the vaccine should be optimally targeted at the elderly, suggests that this finding should hold for many countries with similar age-profiles and age-structured mixing patterns. Ultimately, vaccination remains our only way out of this pandemic, and it is therefore important that the vaccine is deployed as efficiently as possible such that early limited supplies are used to greatest effect.

## Data Availability

For model fitting, we used: data on cases, obtained from the COVID-19 Hospitalisation in England Surveillance System (CHESS) data set that collects detailed data on patients infected with COVID-19; data on COVID-19 deaths, obtained from Public Health England. These data contain confidential information, with public data deposition non-permissible for socioeconomic reasons. The CHESS data resides with the National Health Service (www.nhs.gov.uk) whilst the death data are available from Public Health England (www.phe.gov.uk).

## Author contributions

**Conceptualisation:** Matt J. Keeling.

**Data curation:** Matt J. Keeling; Edward M. Hill.

**Formal analysis:** Sam Moore.

**Investigation:** Sam Moore.

**Methodology:** Sam Moore; Matt J. Keeling.

**Software:** Sam Moore; Matt J. Keeling; Edward M. Hill; Louise Dyson; Michael J. Tildesley.

**Validation:** Sam Moore; Matt J. Keeling; Edward M. Hill; Louise Dyson; Michael J. Tildesley.

**Visualisation:** Sam Moore.

**Writing - original draft:** Sam Moore.

**Writing - review & editing:** Sam Moore; Matt J. Keeling; Edward M. Hill; Louise Dyson; Michael J. Tildesley.

## Ethical considerations

The data were supplied from the CHESS database after anonymisation under strict data protection protocols agreed between the University of Warwick and Public Health England. The ethics of the use of these data for these purposes was agreed by Public Health England with the Government’s SPI-M(O) / SAGE committees.

